# Prevalence use of nonsteroidal anti-inflammatory drugs in the general population with COVID-19 and associated COVID-19 risk, hospitalization, severity, death, and safety outcomes: A systematic review and meta-analysis

**DOI:** 10.1101/2021.05.01.21256428

**Authors:** Huilei Zhao, Shanshan Huang, Kaibo Mei, Wen Shao, Jing Zhang, Jianyong Ma, Wengen Zhu, Yuan Jiang, Peng Yu, Xiao Liu

**Author notes:** Dr. Zhao, Dr. Huang and Dr. Mei are co-first authors. **Corresponding author:** Peng Yu, Xiao Liu.

## Abstract

**Introduction:** Recent reports of potential harmful effects of nonsteroidal anti-inflammatory drugs (NSAIDs) in patients with Corona Virus Disease 2019 (COVID-19) have provoked great concern. Therefore, the safety of NSAIDs is still questioned.

**Methods:** We searched the PubMed, EMBASE, Cochrane Library and Web of Science databases from December 2019 to January 2021 to examine use prevalence for NSAIDs in general, as well as associated COVID-19 risk and outcomes. This study has been registered with PROSPERO (CRD42019132063)

**Results:** We included 25 studies with a total of 101,215 COVID-19 patients. The use of NSAIDs in COVID-19 patients reached 19%. Exposure to NSAIDs was not associated with significantly increased risk of developing COVID-19 (odds ratio [OR]=0.98, 95% confidence interval [CI]: 0.78-1.24; I^2^=82%), hospitalization (OR=1.06, 95%CI: 0.76-1.48; I^2^=81%), mechanical ventilation (OR=0.71, 95%CI: 0.47-1.06; I^2^=38%), and length of hospital stay. Moreover, use of NSAIDs was significantly associated with better outcomes, including severity of COVID-19 (OR=0.79, 95%CI: 71-0.89; I^2^=0%) and death (OR=0.68, 95%CI: 0.52-0.89; I^2^=85%) in patients with COVID-19. Regarding safety outcomes, exposure to NSAIDs was associated with increased risk of stroke (OR=2.32, 95%CI: 1.04-5.2; I^2^=0%), but not with myocardial infraction (OR=1.49; *p*=0.66; I^2^=0%), overt thrombosis (OR=0.76, *p=0*.*50*; I^2^=28%) and major bleeding (*p*=0.61).

**Conclusion:** Based on current evidence, exposure to NSAIDs is not linked to increased odds or exacerbation of COVID-19 in the general COVID-19 population. Furthermore, administration of NSAIDs might have better outcomes and survival benefits in the general COVID-19 population, although potentially increasing the risk of stroke. Use of NSAIDs might be safe and beneficial in COVID-19. Future observational and randomized control trials are needed for further confirmation.

## INTRODUCTION

Corona Virus Disease 2019 (COVID-19) is an acute respiratory disease caused by the novel virus called severe acute respiratory syndrome-associated coronavirus-2 (SARS-CoV-2), which infects cells expressing the angiotensin-converting enzyme (ACE2) receptor and is currently prevalent worldwide[1, 2].

Non-steroidal anti-inflammatory drugs (NSAIDs) such as aspirin, ibuprofen, celecoxib, and indomethacin are inexpensive and readily available drugs widely applied for their antiviral and anti-inflammatory characteristics[3]. Moreover, since most clinical symptoms of COVID-19 usually start from fever, a sign of pain, NSAIDs might be the most commonly used drugs in the general population. Therefore, NSAIDs provoke safety concerns in COVID-19. The concern was initially raised by several cases of COVID-19 reported by French researchers, with worsened symptoms after taking the NSAID ibuprofen[4]. Quite a few reports have revealed various side effects of NSAIDs and opposed their administration in COVID-19 patients[4-6]. Subsequently, the European Medicines Agency and the WHO claimed NSAIDs should not be precluded when clinically indicated, at least not all of NSAIDs, based on limited evidence in the covid-19 epidemic.

NSAIDs exert their effects by suppressing cyclooxygenase enzymes so that the synthesis of prostaglandins (PGs) is reduced. Substances like prostaglandins are closely related to signs of pain, fever, and inflammation in patients with COVID-19[7]. Out of theoretical assumptions, studies suggested that exposure to NSAIDs might increase the risk of COVID-19; moreover, it is speculated that use of NSAIDs in COVID-19 patients may worsen the disease and increase the odds of ICU admission and death[8-11]. By contrast, other articles reported that prior exposure to NSAID does not significantly increase the risk of COVID-19 in general and is even associated with better outcomes (e.g., admission and death) in hospitalized COVID-19 patients, e.g., aspirin[12-14]. In light of the ibuprofen–COVID-19 debate and the rapidly unfolding situation of the current pandemic, this study aimed to 1) assess the prevalence use of NSAIDs in the general COVID-19 population, 2) systematically investigate the associations of NSAID use with the risk of developing COVID-19 and related outcomes, and 3) assess the safety (particularly vascular complications) of NSAIDs in patients with COVID-19.

## METHODS

This study was performed in accordance with PRISMA 2020 guidelines[15] **(Table S1 in Supplemental Materials)** and has been registered on PROSPERO (International prospective register of systematic reviews) (registration number-CRD42019132063).

### Literature search

Two independent researchers (Liu Xiao and Shanshan Huang) searched the PubMed, Cochrane Library, Web of Science, EMBASE, and MedRxiv (https://www.medrxiv.org/) databases from December 2019 to January 2021 for relevant studies without any language restrictions. We performed the search using the following keywords: “Nonsteroidal anti-inflammatory drug” OR “NSAID” AND “2019-novel coronavirus” OR “SARS-CoV-2” OR “COVID-19” OR “2019-nCoV”. Besides, references of related studies were also reviewed to discover more potentially relevant studies. Discrepancy was discussed by two researchers through online meeting until census were reached.

### Study selection/inclusion and exclusion criteria

All included studies should meet the following criteria:1) recorded the use of NSAIDs prior or during the COVID-19 panic of general population. 2) evaluated the association between the use of NSAID and risk of developing COVID-19 and associated outcomes, such as hospitalization, severity and death in COVID-19 patients. Exclusion criteria were: 1) Reports focused on specific population, such as orthopedic and rheumatologic diseases were excluded. 2) Case reports, letters, reviews and expert opinions.

### Data extraction and quality assessment

Two investigators (Xiao Liu and Shanshan Huang) independently extracted following information: name of first author’s name, publication year, country, study design, sample size, mean or median age, sex, measure of exposure to NASIDs. Methods of astern COVID-19. In present study, the intensive care unit (ICU) admission was also considered as severe COVID-19 event. The quality and risk of bias of included studies reporting use of NSAIDs and associated outcomes (e.g., risk of COVID-19, severe COVID-19, and death) was assessed by the Joanna Briggs Institute (JBI) critical appraisal check-list and Newcastle–Ottawa quality scale (NOS), respectively[16, 17]. [18]We regarded studies with an NOS of ≥6 stars as medium to high-quality studies; otherwise, are considered low-quality.

### Outcomes

We firstly assessed the incidence of use of NASIDs in COVID-19 patients. Then the association between NASIDs and the risk of developing COVID-19 infection and associated outcomes (e.g., mechanical ventilation, hospitalization, severe COVID-19 infection, and mortality) was assessed. Finally, we focused on the safety of NASIDs use in COVID-19, particular vascular complications, such as bleeding, myocardial infraction, and stroke.

### Statistical analysis

Meta-analysis was performed using international STATA15.0 (StataCorp, College Station, Texas) and RevMan5.3 (Review Manager [RevMan], version5.3, Cochrane Collaboration) software. The OR and 95% confidence intervals (95%CIs) were taken to evaluate the merged results. Risk ratio and hazard ratio from other studies were regarded as OR. If not available, ORs could be obtained through calculating events and total numbers of patients in two groups. If both unadjusted and adjusted RRs existed in one study, we extracted both and pooled the unadjusted and adjusted, respectively. If multi-adjusted ORs were reported, we used the most completely adjusted one. The calculation and aggregation of natural logarithm of the OR (log [OR]) and its standard error (SElog [OR]) were conducted by Revman 5.3. As for evaluation of heterogeneity, Cochran’s chi-square test and I^2^ statistic was used. The random effects model were used to pooled the results. We also performed a subgroup analysis of types of NSAIDs which based on studies adjusted for confounding. The evaluation of publication bias was conducted by Egger’s test and Begg’s funnel plot. *P* < 0.05 represent there is statistical significance. Sensitivity analysis was conducted by omitting one study each.

## RESULTS

### Study selection

After searching the PubMed, Cochrane Library, EMBASE, and MedRxiv databases, we obtained 573 potentially relevant articles. Of these, 63 were duplicates. In addition, 447 studies were excluded after examining their titles and abstracts. Of the remaining 53 articles, 28 were excluded after a detailed review of the full text for the following reasons_:_1) 8 reports assessed other respiratory infections; 2) 7 were letters, reviews, or case reports; 3) 6 studies assessed suspected COVID-19 cases instead of confirmed disease; 4) 4 articles were based on other diseases (e.g., rheumatologic diseases, cancers, and coronary artery diseases); 5) 3 reports were based on duplicated populations. Finally, 25 publications (23 cohorts) [11-13, 19-40] were included in the present analysis **(Figure 1)**.

**Figure 1.**
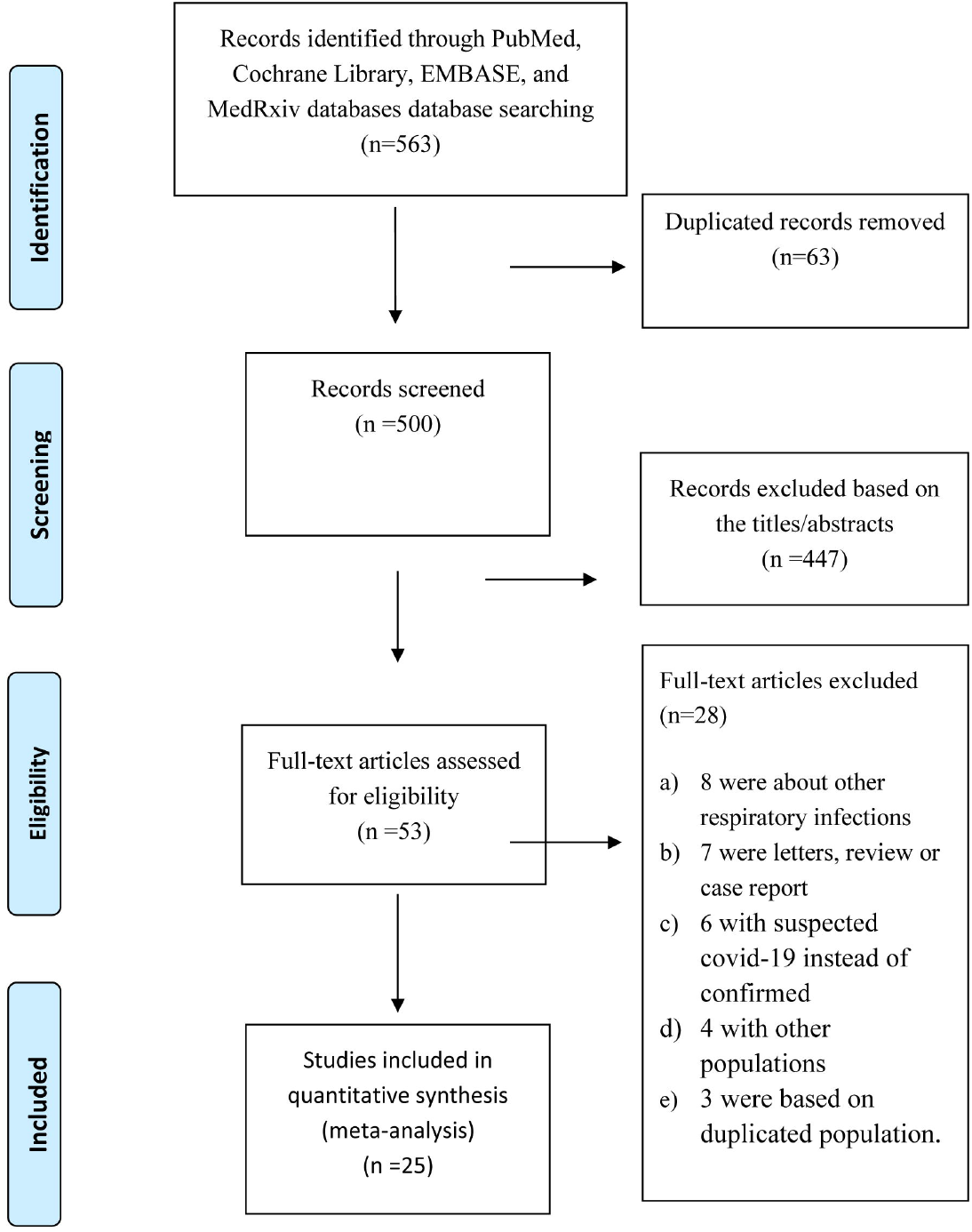
Flow chart of study selection.

### Study characteristics and quality

The basic characteristics of the included studies are shown in Table 1. All the included patients were general COVI-19 cases. The mean age ranged from 64 to 67 years. The doses of NSAIDs were not reported in almost all studies, except one [38] in which 81 mg aspirin was administered. The diagnosis of COVI-19 was confirmed by SARS-COV-2 PCR in most of the studies; one[40] used ICD-10, and one[13] combined clinical and SARS-COV-2 PCR findings. The majority of studies were retrospective or prospective cohort observational trials; besides, 3 trials[22, 24, 29] were case-control studies. The assessment results showed that all articles were of medium to high quality **(Table S2-3 in Supplemental Materials)**.

**Table 1.**
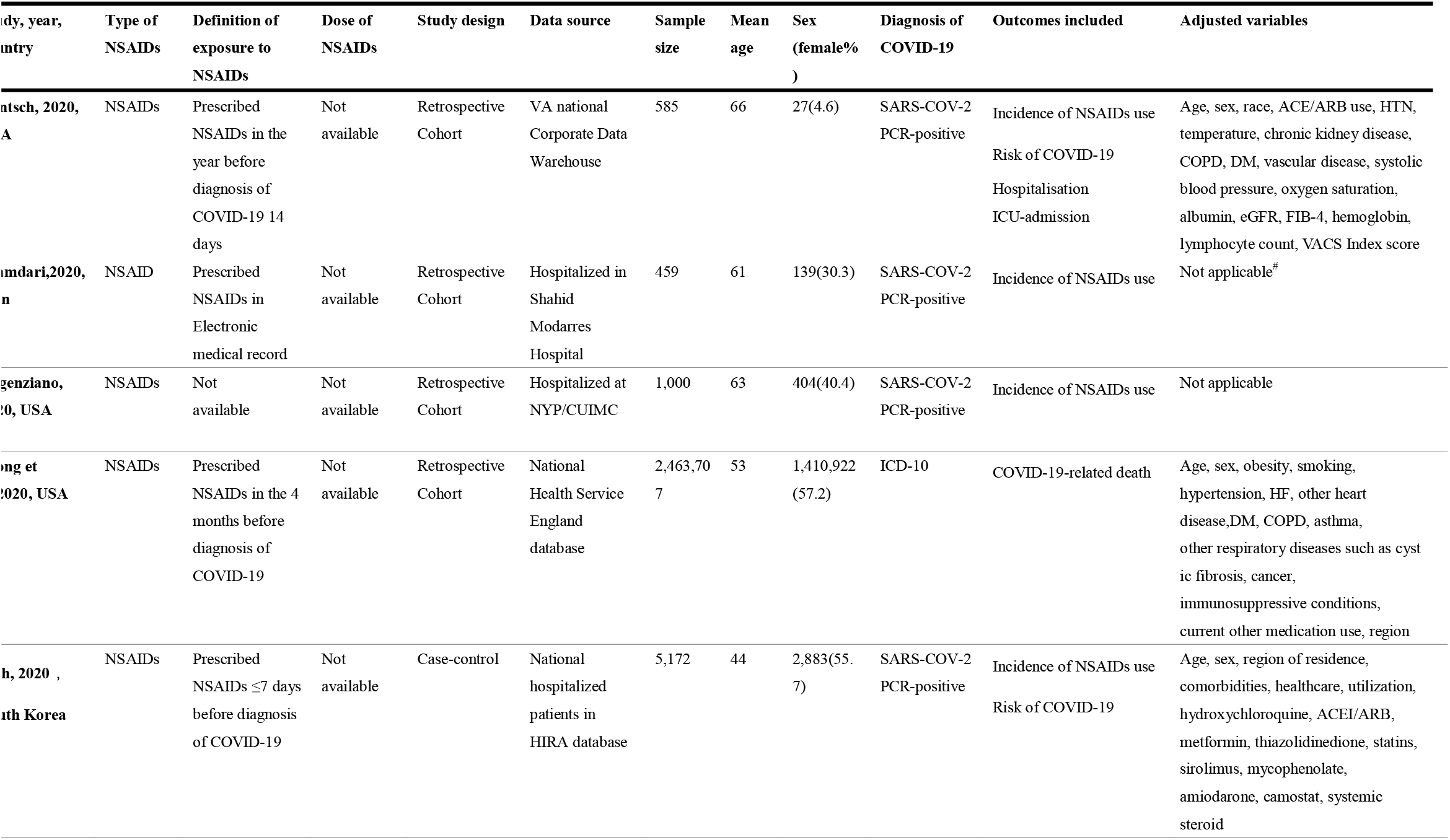
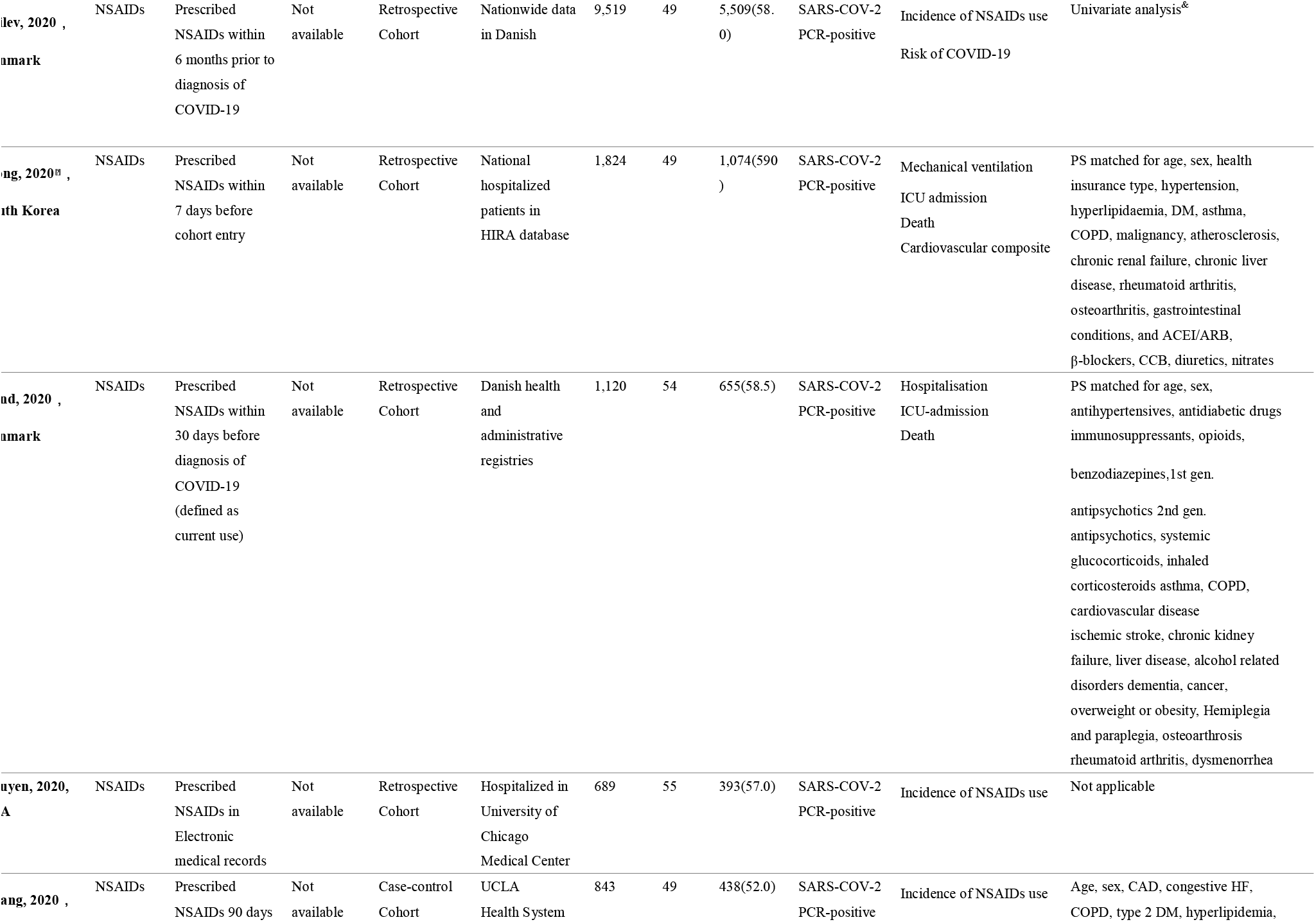
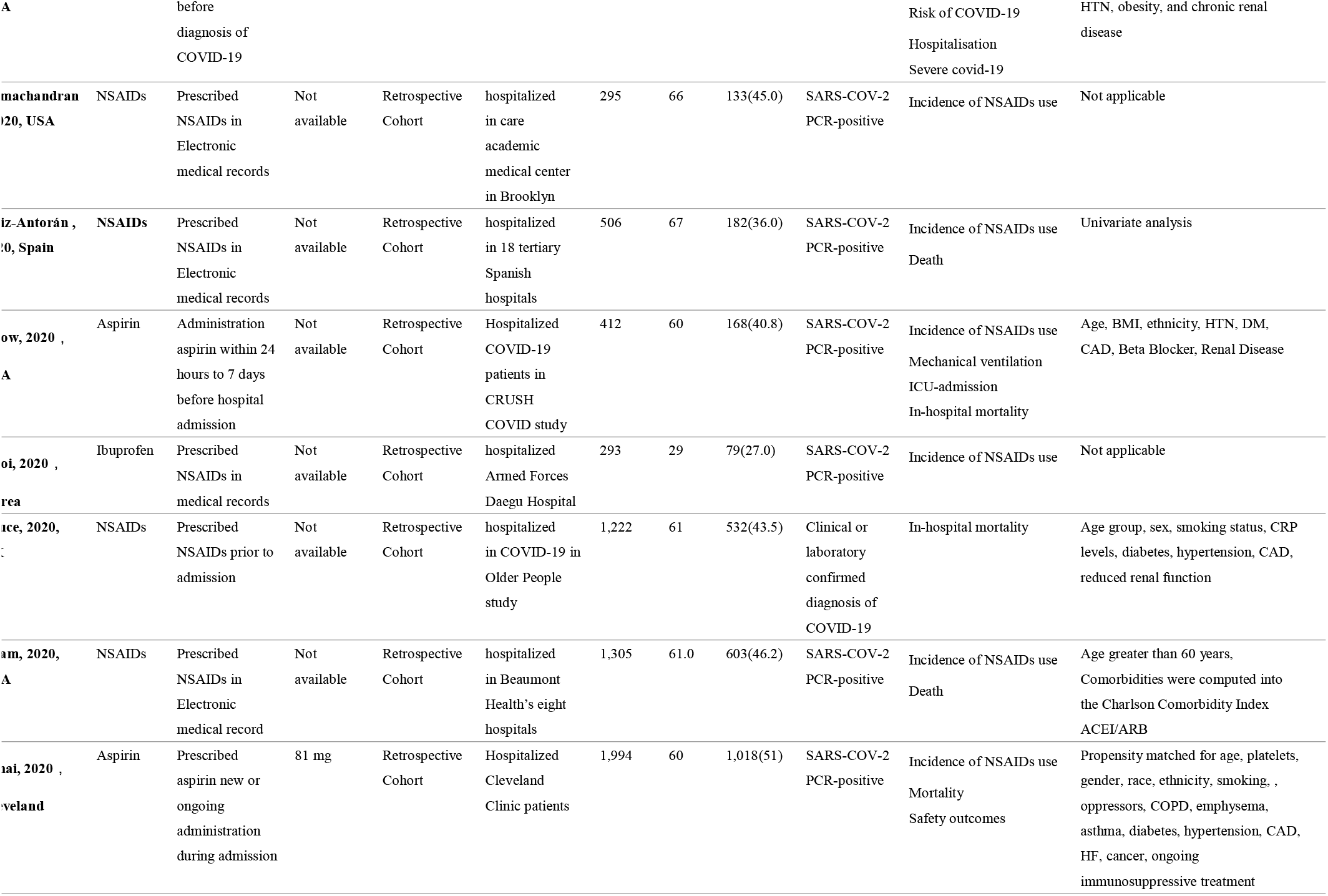
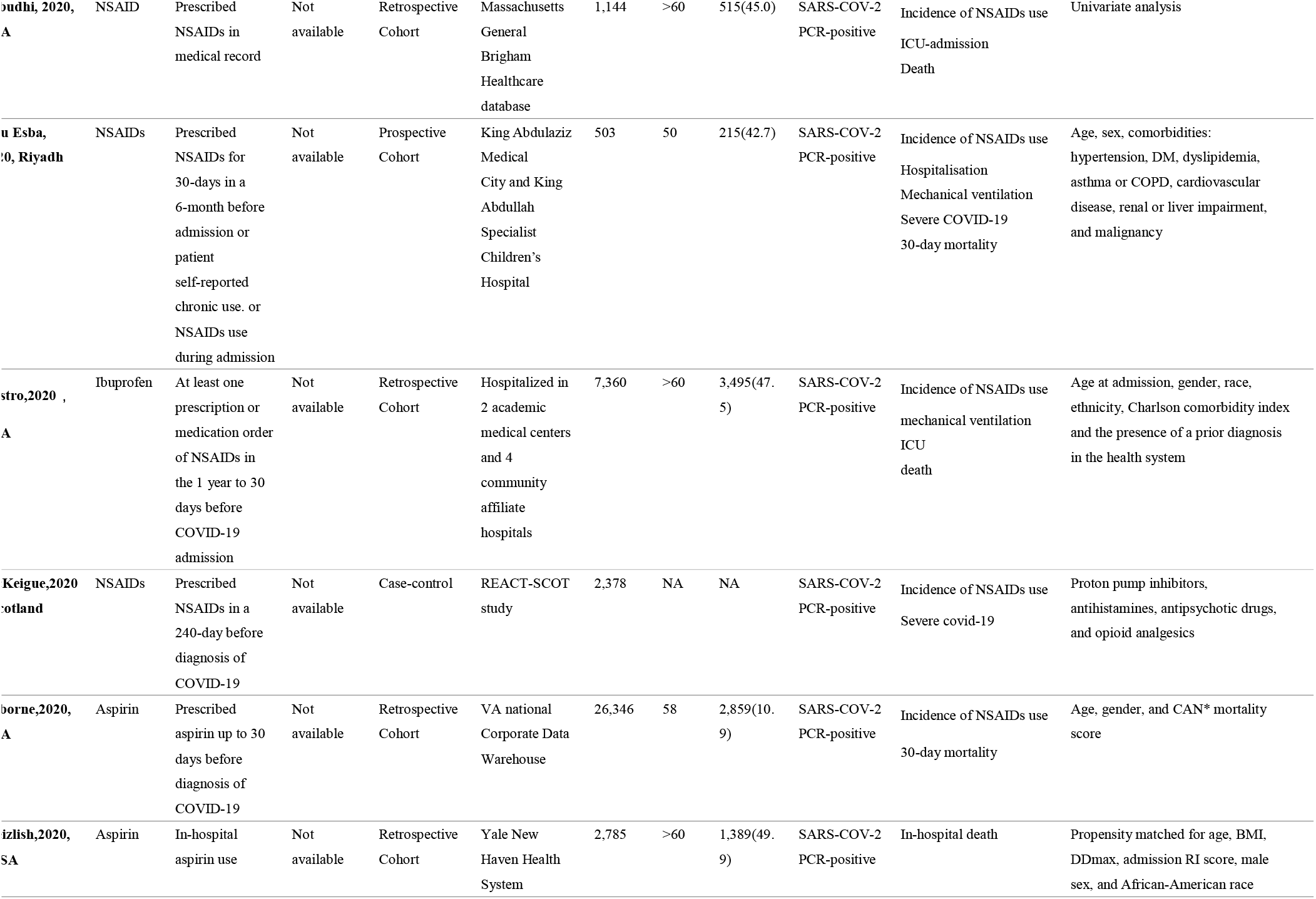
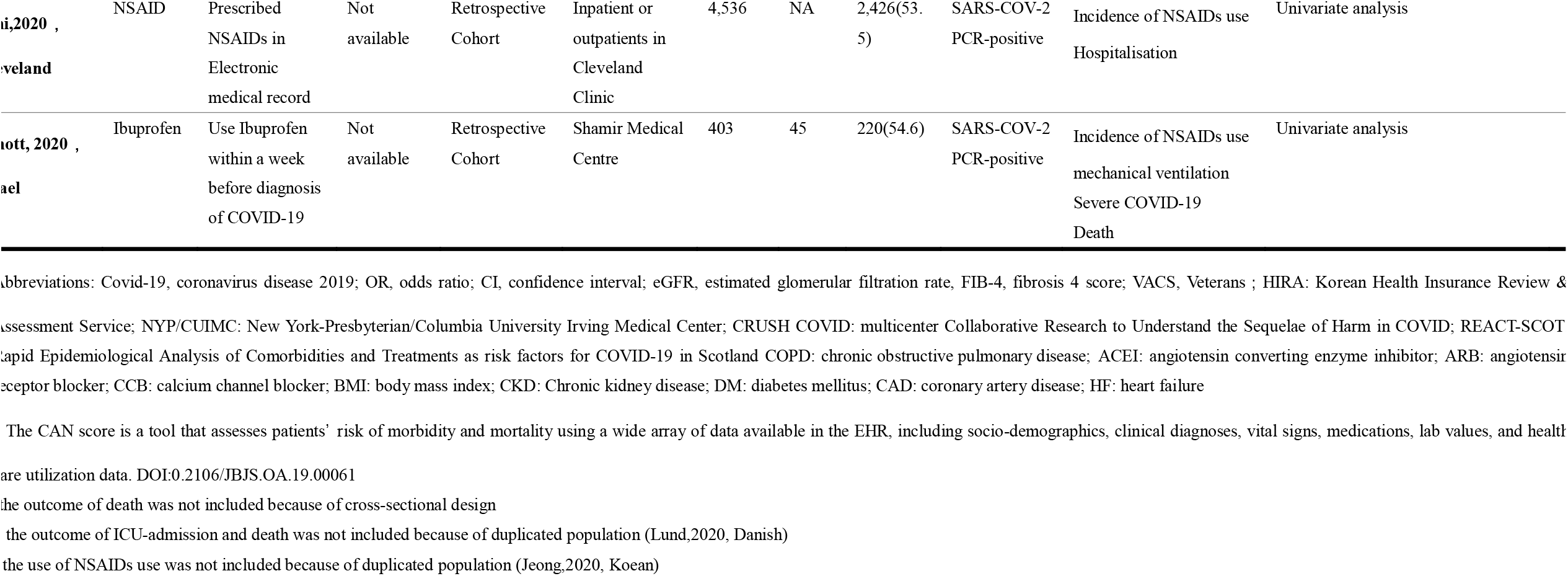
Basic information of included studies.

### Use prevalence for NSAIDs in patients with COVID-19

Twenty-two articles with 106,421 patients[8-13, 19-26, 29-34, 36, 38, 39, 41-49] reported the use of NSAIDs in general COVID-19 cases. The pooled results showed a use prevalence for NSAIDs in COVID-19 patients reaching 19% (95%CI: 14%-23%), with significant heterogeneity (I^2^=99%) **(Figure 2a)**. Subgroup analysis showed no statistical differences in NSAID use prevalence across age, study designs, regions, and sample sizes **(Figure S1a-d in Supplemental Materials)**.

**Figure 2.**
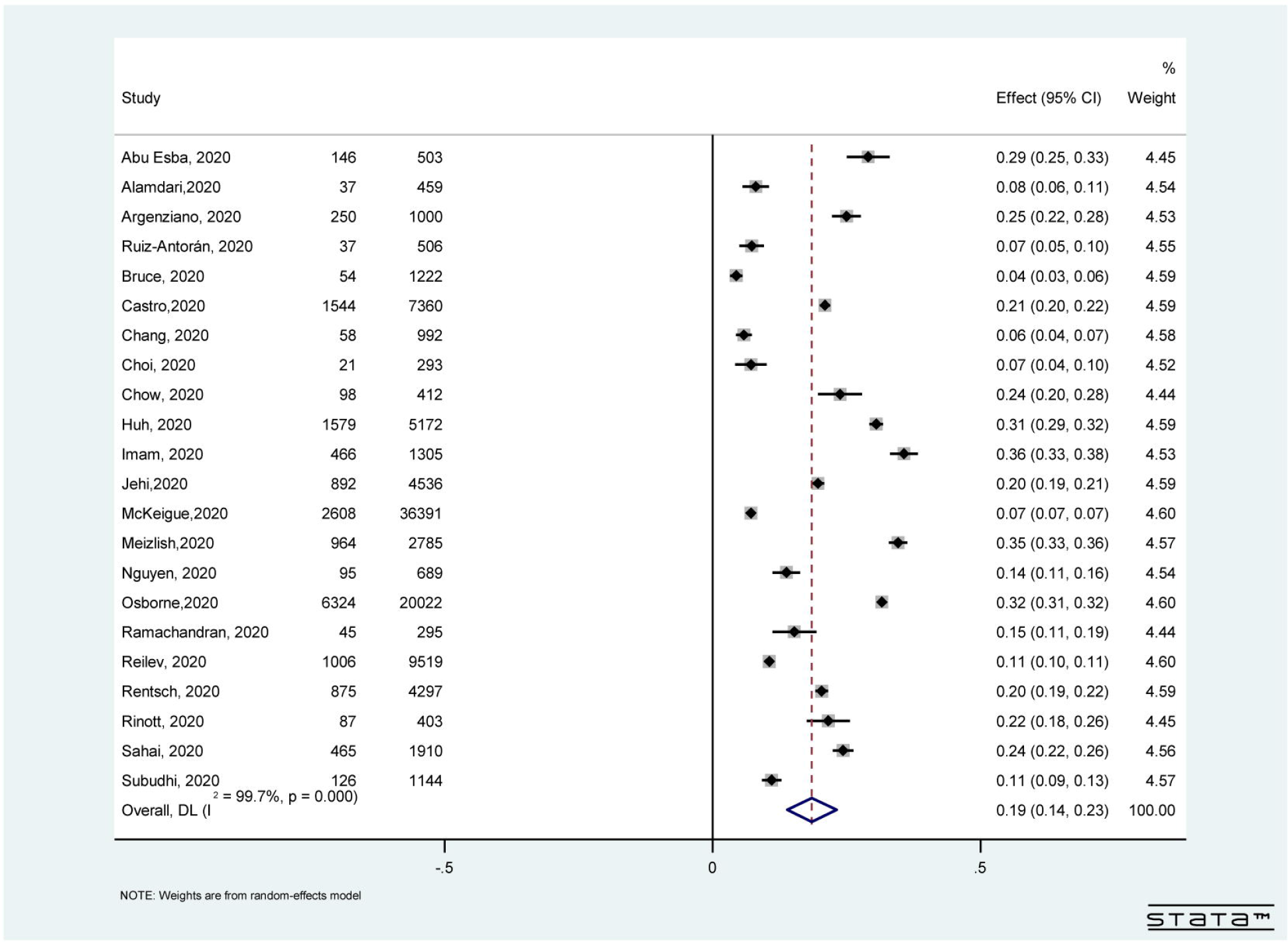
Forest plot for the prevalence use of **nonsteroidal anti-inflammatory drugs** in general COVID-19 population.

### The impact of NSAIDs on the risk of COVID-19

Five studies[8, 9, 22, 42, 45] with 17,239 cases among 343,286 individuals reported the effect of NSAID use on the risk of COVID-19 infection. The pooled analysis of raw data showed that use of NSAIDs did not increase the risk of COVID-19 (crude OR=0.93, 95%CI: 0.78–1.12; I^2^=83%) **(Figure 3a)**. Three studies[8, 22, 42] evaluated the association between NSAIDs and COVID-19 infection by multivariate analysis. Consistently, there was no significant association between NSAID use and COVID-19 risk (adjusted OR=0.93, 95%CI: 0.82–1.06; I^2^=34%) **(Figure 3b)**.

**Figure 3.**
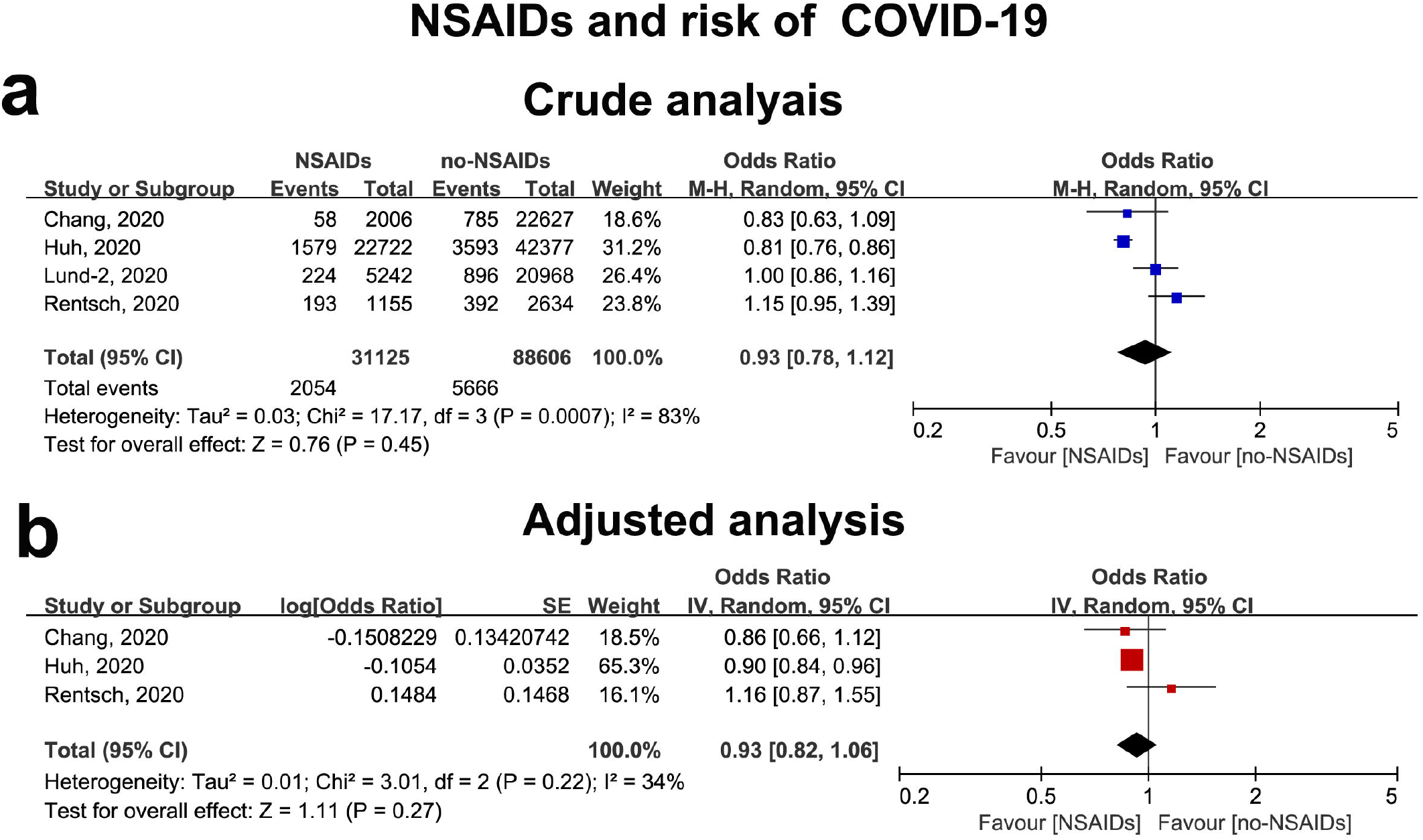
Forest plot for the association between nonsteroidal anti-inflammatory drugs and risk of COVID-19 infection. **A**. Crude effect size of the association between nonsteroidal anti-inflammatory drugs and risk of COVID-19 infection. **B**. Adjusted effect size of the association between nonsteroidal anti-inflammatory drugs and risk of COVID-19 infection.

### The impact of NSAID use on hospitalization in patients with COVID-19

There were 9 publications[11, 21, 22, 26, 28, 31, 35, 36, 50] with 10,955 cases/30,921 COVID-19 patients included in the analysis of NSAIDs and hospitalization. The pooled results of the unadjusted analysis showed that use of NSAIDs did not significantly elevate the risk of hospital admission (crude OR=1.05, 95%CI: 0.63–1.73; I^2^=97%) **(Figure 4a)**. Three studies further reported the adjusted results, with a summary OR of 1.06 (95%CI: 0.76–1.48; I^2^=81%) **(Figure 4b)**. Subgroup analysis showed all types of NSAIDs did not significantly increase the risk of severe COVID-19 after adjustment (ibuprofen, OR=0.82, 95%CI: 0.54–1.23; naproxen, OR=0.66, 95%CI: 0.50–0.87) **(Figure S2 in Supplemental Materials)**.

**Figure 4.**
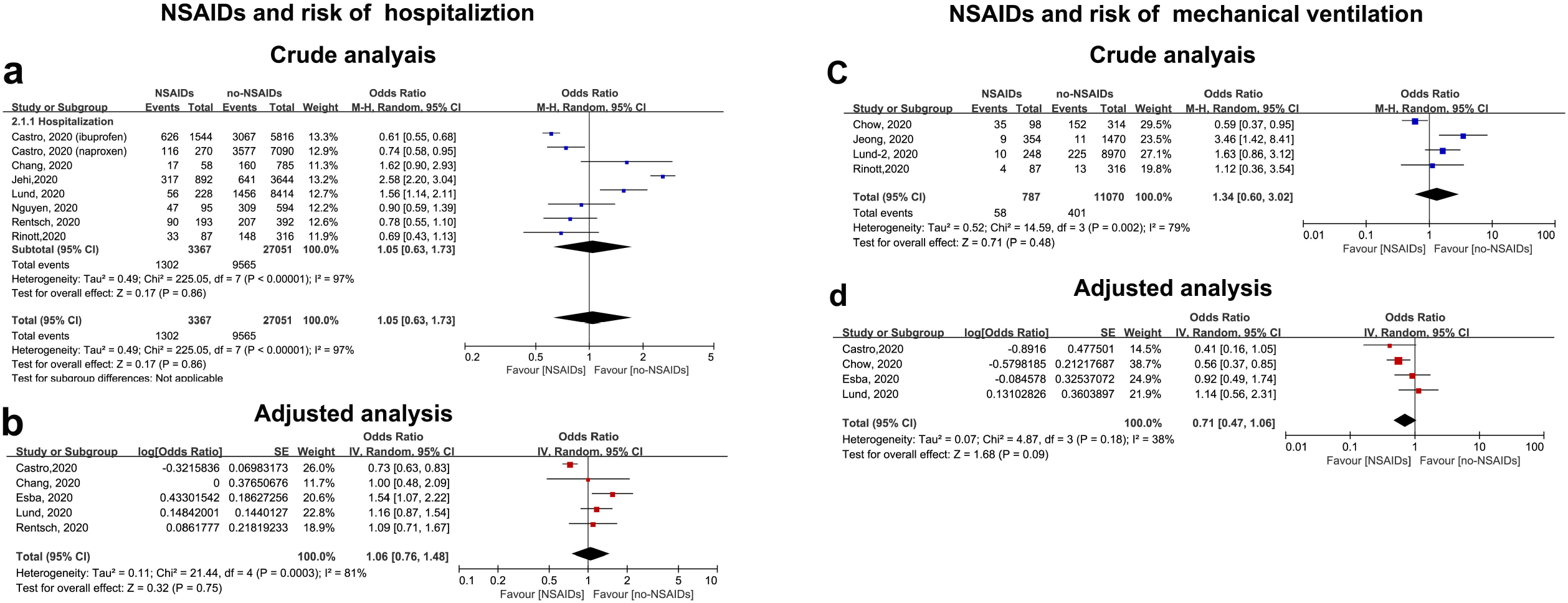
Forest plot for the association between nonsteroidal anti-inflammatory drugs and hospitalization, mechanical ventilation in COVID-19 patients. **A**. Crude effect size of the association between anti-inflammatory drugs and hospitalization in COVID-19 patients. **B**. Adjusted effect size of the association between anti-inflammatory drugs and mechanical ventilation in COVID-19 patients.

### The impact of NSAID use on length of hospitalization in patients with COVID-19

There were 3 publications that reported the hospitalization length based on NSAID use **(Table 2)**. We did not pool the results due to sufficient data. Jeong et al.[51] reported a median length of hospitalization of 12 days among NSAID users versus 13 days among non-users in COVID-19 patients. Another retrospective cohort including 412 American COVID-19 patients reported that there was no significant difference in the length of hospital stay between aspirin use and no-aspirin use[12]. A prospective cohort study that included 503 patients in Asia also showed that acute or chronic use of NSAID did not increase the length of hospital stay compared to non-NSAID users[11] (*p*=0.63). Collectively, current evidence showed NSAIDs did not increase the length of hospitalization in patients with COVID-19.

**Table 2.** Studies reported the relationship between use of NSAIDs and length of hospitalized days in patients with COVID-19.

| References<br>(first author, year, country/region) | Study participants | Study design | Sample size | Length of hospitalization |  |  |
| --- | --- | --- | --- | --- | --- | --- |
|  |  |  |  | Values (Median (IQR), days) |  |  |
|  |  |  |  | Group1 | Group2 | *P* |
|  |  |  |  | NSAID users | Non-users |  |
| Jeong,2020, South Korea | National hospitalized COVID-19 patients in HIRA database | Retrospective Cohort | 1,824 | 12 | 13 | Na |
| Chow,2020, USA | Hospitalized COVID-19 patients in CRUSH COVID study | Retrospective Cohort | 412 | No Aspirin<br>8 (3-19) | Aspirin<br>9 (5-17) | 0.91 |
| Abu Esba,2020, Riyadh | National COVID-19 hospitalized patients in Health Affairs | Prospective Cohort | 503 | NSAID users<br>7 | Non-users<br>8 | 0.63 |

\*RUSH COVID: multicenter Collaborative Research to Understand the Sequelae of Harm in COVID; COVID-19: Corona Virus Disease 2019; HIRA: Korean Health Insurance Review & Assessment Service;

### The impact of NSAID use on mechanical ventilation in patients with COVID-19

There were 6 studies[11, 12, 21, 27, 28, 36] with 459 cases/11,857 patients using mechanical ventilation. Both the pooled results of crude (crude OR=1.34, 95%CI: 0.60–3.02; I^2^=79%) and adjusted analysis showed no significant risk of mechanical ventilation with the use of NSAIDs in COVID-19 cases (adjusted OR=0.71; 95%CI: 0.47–1.06; I^2^=38%) **(Figure 4c-d)**. Subgroup analysis showed all types of NASIDs did not significantly increase the risk of severe COVID-19 after adjustment (aspirin, OR=0.56, 95%CI: 0.47–1.06; ibuprofen, OR=0.75, 95%CI: 0.22–2.50; naproxen, OR=0.36, 95%CI: 0.03–3.96) **(Figure S3 in Supplemental Materials)**.

### The impact of NSAID use on severe COVID-19 infection

Twelve articles involving 6,284 severe COVID-19 cases among 69,942 patients[8-12, 20-22, 27-29, 36, 39, 43-45, 49] reported the association between NSAID use and COVID-19 severity. As shown in **Figure 5a**, treatment with NASIDs was not significantly associated with the likelihood of severity in COVID-19 (crude OR=1.01, 95%CI: 0.73–1.24; I^2^=85%). Furthermore, pooled OR in multivariate analysis[8, 9, 12] showed significantly reduced risk of severe COVID-19 infection (adjusted OR=0.79, 95%CI: 0.71–0.89; I^2^=0%) **(Figure 5b)**. Subgroup analysis showed all types of NASIDs did not significantly increase the risk of severe COVID-19 after adjustment (aspirin, OR=0.57, 95%CI: 0.38–0.86; ibuprofen, OR=0.98, 95%CI: 0.59–1.65; naproxen, OR=0.79, 95%CI; 0.52–1.20) **(Figure S4 in Supplemental Materials)**.

**Figure 5.**
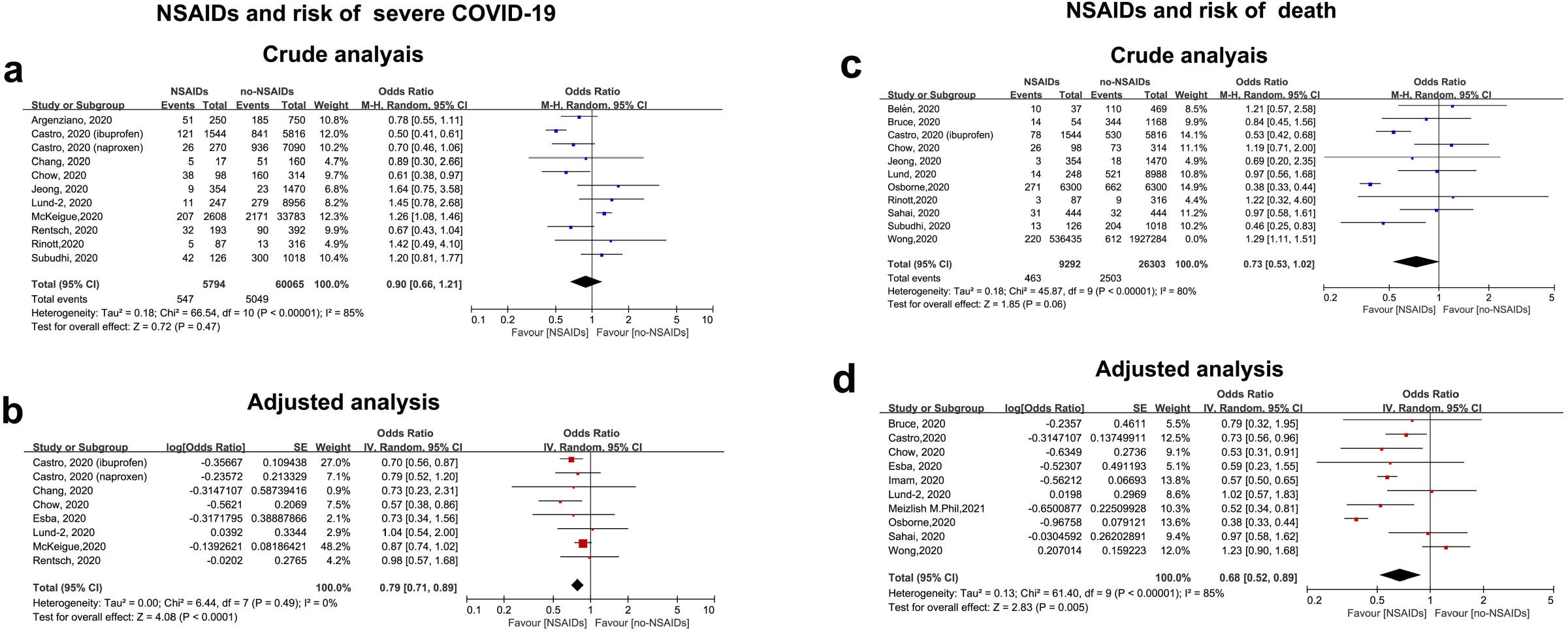
Forest plot for the association between nonsteroidal anti-inflammatory drugs and severity, death in COVID-19 patients. **A**. Crude effect size of the association between anti-inflammatory drugs and severity in COVID-19 patients. **B**. Adjusted effect size of the association between anti-inflammatory drugs and death in COVID-19 patients.

### The impact of NSAID use on all-cause death in patients with COVID-19

We included fifteen studies[11-13, 19, 21, 25, 27, 28, 36, 52] with 2,966 cases among 35,595 patients in the analysis of death and NSAID use in COVID-19. We observed a decreased but non-significant risk of death after NSAID use and in COVID-19 patients (crude OR=0.73, 95%CI: 0.53–1.02; I^2^=80%) **(Figure 5c)**.

Notably, the pooled results of adjusted analysis showed that use of NSAIDs significantly decreased the risk of death in COVID-19 (adjusted OR=0.68, 95%CI: 0.52–0.89; I^2^=85%) (Figure 5d). Subgroup analysis showed all types of NASIDs did not significantly increase the risk of COVID-19 death after adjustment (aspirin, OR=0.41, 95%CI: 0.22–2.50; ibuprofen, OR=0.83, 95%CI: 0.69-0.99; naproxen, OR=0.92, 95%CI: 0.72–1.17) **(Figure S5 in Supplemental Materials)**.

### Safety of NSAID use in patients with COVID-19

The cardiovascular adverse effects of NSAIDs, such as major bleeding, heart failure, and major coronary events, are well established in the general population[53]. Therefore, we also focused on the potentially harmful effect of NSAIDs on vascular complications in COVID-19. Two propensity score matched studies reported NSAID-related safety outcomes (myocardial infarction, stroke and major bleeding)[27, 38]. The pooled results showed prescription of NSAIDs significantly increased the risk of stroke (PS matched OR=2.06, 95%CI: 1.04–5.2; p=0.16; I^2^=0%) **(Figure 6a)**, but not significantly increased myocardial infraction (propensity score matched OR=1.48, 95%CI: 0.25–8.92; I^2^=0%) and overt thrombosis (OR=0.76, 95%CI: 0.34–1.70; I^2^=28%)**(Figure 6b-c)**. Moreover, Chow et al.[12] found no significant difference (6.1% aspirin vs. 7.6% non-aspirin, *p*=0.61) in major bleeding between groups in the crude analysis.

**Figure 6.**
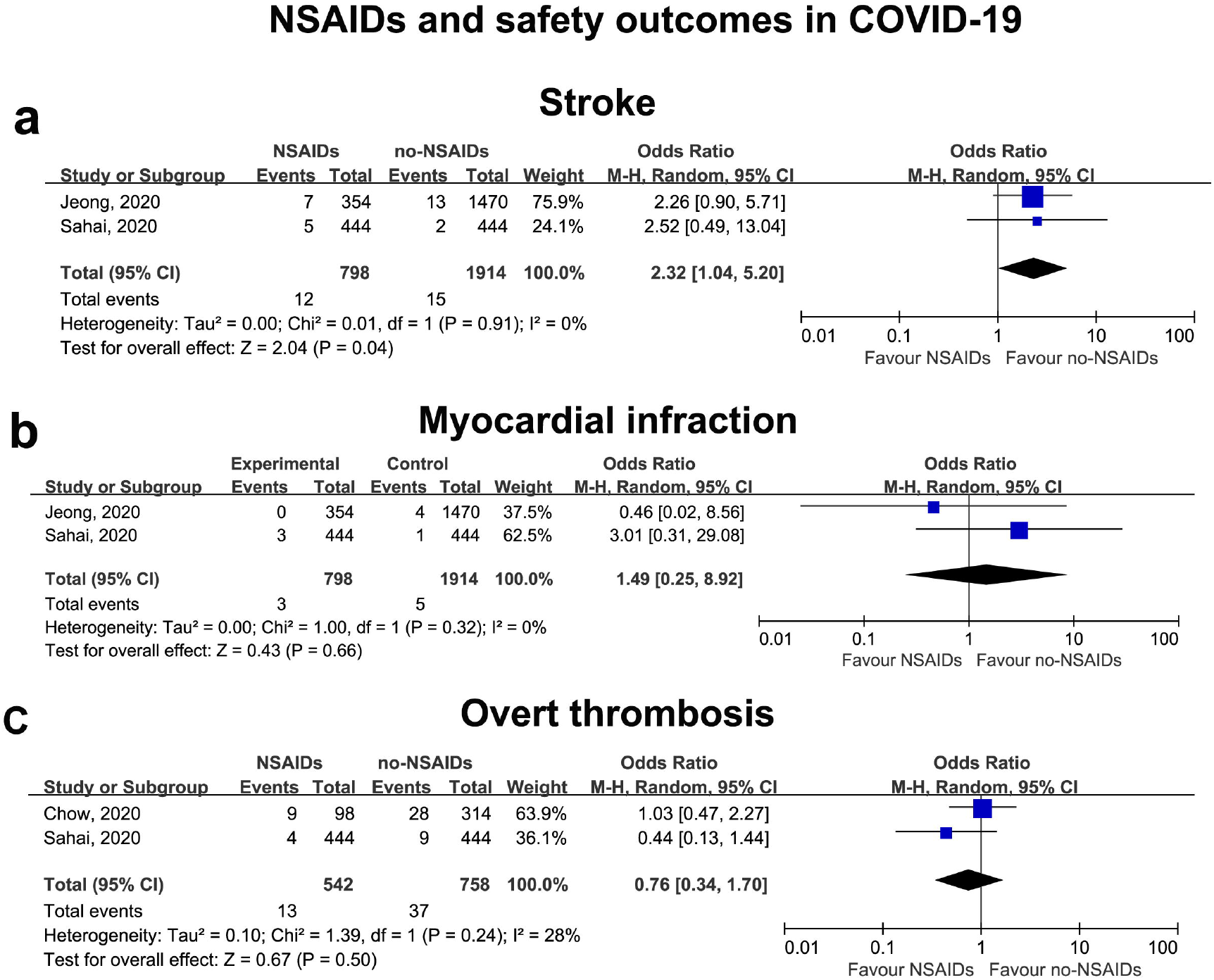
Forest plot for the association between nonsteroidal anti-inflammatory drugs and safety outcomes in COVID-19 patients. **A**.Stroke. **B**. Myocardial infraction. **C:** Over thrombosis

### Publication bias and sensitive analysis

No publication bias for susceptibility to COVID-19 was detected by the funnel plot or the Egger’s test (*p*>0.1), although these tests are not recommended for outcomes for which few studies are included (N<10) **(Figure S6 in Supplemental Materials)**.

## DISCUSSION

At present, no consensual guidelines are available regarding NSAIDs use in COVID-19, reflecting a paucity of data in this regard. To the best of our knowledge, this study is the first meta-analysis that comprehensively assessed the incidence of NSAID use and associated outcomes in the general COVID-19 population. Our results showed a prevalence of NSAID use reaching 17% among the general population with COVID-19, which suggested the use of NSAIDs in patients with COVID-19 is not uncommon. Furthermore, based on current evidence, we found exposure to NSAIDs, including aspirin, ibuprofen and naproxen, was not associated with increased risk of developing COVID-19 or worse outcomes in patients with COVID-19. Notably, a significantly increased risk of thrombotic stroke was found in patients with COVID-19 administered NSAIDs (OR=2.32, *p*<0.04).

There is sound evidence that SARS-CoV-2 spike glycoprotein directly binds to the host cell’s ACE2 receptor, which is highly expressed in human lung tissue, gastrointestinal tract, and arterial smooth muscle cells *in vivo*[54]. Indeed, the ACE receptor in humans plays an important role in the pathophysiology of SARS-CoV-2 infection*[55, 56]*. Several drugs with the potential of increasing ACE2 expression have provoked great concern for potentially contributing to the spread of COVID-19 in the population, including ACEI/ARB[57] and NSAIDs[4]. However, as suggested by some researchers, there is a long way from bench to bedside. The evidence derived from mechanistic or theoretical pharmacology should be cautiously interpreted when drawing conclusions[58]. A number of examples can be found in the literature where evidence from mechanistic studies does not always corroborate data from clinical trials. For example, studies have shown that co-administration of ibuprofen and aspirin can counteract the antiplatelet effectiveness of aspirin as detected by thromboxane levels[59]. This hypothesis, however, was refuted in a large randomized controlled trial[60]. In a previously report, we demonstrated that ACEI/ARB is not associated with increased risk of COVID-19 infection and worse outcomes in the general population or hypertensive patients[61]. Similarly, regarding NSAIDs, a potential risk increase in COVID-19 with the use of NSAIDs was not found. There are several possible explanations for these findings. Firstly, although animal studies showed that ACE2 receptor expression is significantly increased by ibuprofen[62], a recent report found two commonly used NSAIDs, including ibuprofen and meloxicam, have no effect on ACE2 expression, viral entry, and viral replication in a mouse model of SARS-CoV-2 infection[63]. Furthermore, no human studies have assessed the effect of ibuprofen on ACE2 expression, particular in the lung tissue. Secondly, whether there is a positive dose-response relationship between ACE2 expression and the risk of COVID-19 infection remains unknown. The most recent evidence derives from COVID-19 and inflammatory bowel disease. Higher ACE2 protein expression was shown in terminal ileum and colon in patients with inflammatory bowel disease compared with controls[64]. However, there is currently no evidence of increased risk or aggravated outcomes in patients with inflammatory bowel disease in the context of COVID-19[65]. Collectively, based on current data, both experiment and clinical, we suggest that there is no evidence of a positive association between NSAIDs and the risk of COVID-19.

Recent reports of potential harm (e.g., worsening symptoms) by NSAIDs in patients with COVID-19 have attracted widespread attention, and several medical agencies still have inconsistent opinions based on limited evidence[4]. In the present study, we also found the use of NSAIDs had a potential benefit in COVID-19. Indeed, NSAID use reduced the incidence of severe COVID-19 infection and all-cause mortality after adjustments. These results corroborated our previous study of ACEI/ARB, which also evoked great concern based on similar reasons regarding COVID-19[61]. There are several potential interpretations. Firstly, although some literature reviews and investigators recommend to avoid NSAIDs based on previous studies showing negative outcomes in ibuprofen users in the treatment of acute respiratory tract infections, these results might be affected by selection bias[66, 67]. The NSAID groups might have higher disease severity compared with nonusers. For example, whilst Joeng et al.[27] found an increased composite outcome (in-hospital death, ICU admission, mechanical ventilation use, or sepsis) in the NSAID group, elevated event rate was not found compared with individuals administered paracetamol, a drug used for similar indications as NSAIDs[27]. Furthermore, it is thought that the pathophysiology and transmission of COVID-19 show differences in behavior even from other respiratory diseases[68]; therefore, the harmful effects of NSAIDs in individuals infected by other respiratory viruses might not be generalized to COVID-19. In contrast, a recent propensity score matched study of 7747 individuals showed that use of NSAIDs is not associated with 30-day intensive care unit admission or death in patients hospitalized with influenza[69]. Secondly, NSAIDs are well-known anti-inflammatory drugs that inhibit the cyclooxygenase (COX) isoforms COX-1 and COX-2. It is well-established that hyper-inflammatory responses underlie the pathology of severe COVID-19. Indeed, cytokine production is significantly increased in COVID-19. Recent findings demonstrated that NSAIDs decreased the production of a subset of proinflammatory cytokines, including Interleukin 6, and tumor necrosis factor alpha [70, 71], which might reduce the incidence of cytokine storm in COVID-19[70, 71].

Our results might be generalizable to a diverse population. In accordance with the above findings, several cohort studies also found no outcome worsening in COVID-19 patients with coexisting comorbidities, including rheumatic disease[72], cancer[73], and coronary artery diseases[74]. For example, a national cohort study based on 1708 781 individuals with rheumatoid arthritis/osteoarthritis showed that the current use of NSAIDs is not associated with a reduced risk of COVID-19 related death after adjustment[72]. Furthermore, another cohort with a small sample size (N=183) in China found that pre-hospitalization use of low-dose aspirin is not associated with mortality in patients with CAD hospitalized with COVID-19 infection (OR=0.944, *p*=0.89)[74]. Collectively, these results showed treatment with NASIDs is also safe for other patient populations, and individuals taking NASIDs for secondary prevention should continue their treatment.

A previous study found NSAIDs use, compared with non-use, is associated with increased risk of ischemic stroke in patients with acute respiratory infections[75]. Consistently, we also found a significantly increased risk of stroke, but no overt thrombosis or myocardial infarction rate elevation after NSAID use in hospitalized COVID-19 patients by pooling two propensity score-matched studies. The somewhat contradictory results were interesting. A possible explanation between thrombosis and clinical findings is the examination of overt thrombosis and microthrombosis, which is usually the cause of thrombosis stroke. It is well-known that the presence of microthrombi does not correlate with overt thrombosis. Microthrombosis is better diagnosed with video-microscopes, dark-field images, and spectral images. Therefore, further researches should use more effective tools to comprehensively elucidate the effect of aspirin or other NSAIDs on thrombosis.

It is well-known that NSAIDs increase the risk of major bleeding, e.g., upper gastrointestinal complications. Chow et al.[12] found no significant increase in major bleeding in patients administered aspirin, which might be explained by the fact that patients with COVID-19 are frequently hypercoagulable, and thrombocytopenia is uncommon in COVID-19 patients. However, considering the observational study design and limited sample size, ongoing COVID-19 clinical trials involving aspirin or NSAIDs would be helpful in providing sounder evidence on safety outcomes.

Age is an important confounding factor that might influence our results. Elderly people with comorbidities are more prone to routinely take NSAIDs[76] Although most of our data were adjusted for age, we cannot fully exclude the potential age-related bias. However, the NSAID groups were older in most of the included studies, and age-related adverse events cannot be underestimated. Furthermore, Wong et al. [40] reported that there is no association between NSAID use and COVID-19 related-death both in young and older individuals, and the estimated effect did not differ by age in all adjustment models. In general, current evidence suggested there is no interaction between age and NSAIDs in COVID-19.

Some authors suggested that different types of NSAIDs might lead to variable capabilities of suppressing the enzymatic activities of COX-1 and/or COX-2, affecting the production profile of downstream eicosanoids, resulting in a variety of effects[5, 6]. In the above subgroup analysis, we found that all types of NSAIDs (aspirin, ibuprofen, and naproxen) were safe for the primary care of all COVID-19 patients, which is consistent with our main results. However, because only a limited study categorized the types of NSAIDs, more research is needed to confirm our results.

### Study strengths and limitations

The greatest strength of this study was its large sample size, and all trials were general population-based. We examined the use rate of NSAIDs in COVID-19 cases and comprehensively assessed the related outcomes, including the risk of COVID-19, hospital admission, severity, mechanical ventilation and death, as well as safety outcomes. As readily available and inexpensive drugs, these results revealed benefits for the clinical use of NSAIDs during the COVID-19 pandemic.

We recognize possible limitations as well. 1) There are intrinsic limitations associated with any observational study, which is unable to prove a causal relationship. 2) The included individuals were adult, and the effects of NSAIDs in young individuals and children should be further studied. 3) Measured and unmeasured factors, such as various underlying diseases, may have influenced the above results. For example, it is thought that there is a great gender difference in COVID-19. However, the limited data prevented sex-specific subgroup analysis. 4) Some data, including age and specific drug doses, were incomplete in most included studies and could not be further analyzed. However, Wong et al.[40] reported that high or low ibuprofen or naproxen is not linked to increased risk of COVID-19 death in the general population, which suggests NSAID use is safe at common clinical doses. 5) Significant heterogeneity was observed across the included studies, which might result from between-study differences, including differences in exposure to NSAIDs, types of NSAIDs and analytical strategies. Finally, well-randomized controlled clinical trials are still required for further assessment of the clinical benefit or harm of NSAIDs at proper doses in the prevention and treatment of COVID-19, while also trying to avoid their known side effects.

## Conclusions

The current findings support the notion that exposure to NSAIDs is not linked to an elevated likelihood of susceptibility to or exacerbation of COVID-19; in addition, administration of NSAIDs might be beneficial to COVID-19 outcomes to some extent. Thus, proper use of NSAIDs in COVID-19 is recommended, rather than absolute rejection. It is worth pointing out that this analysis was mostly based on observational studies, and ongoing trials (NCT0432563339, NCT0438276840, NCT0433462941 and NCT04344457) are expected to demonstrate the role of NSAIDs in the management of COVID-19. What’s more, future investigations should not only focus on the use of NSAIDs as a therapeutic option but also continue to examine the effects of pre-admission NSAID use on the risk of COVID-19 as well as COVID-19 outcomes and mortality in the general population.

## Data Availability

All data generated or analyzed during this study are included in this published article [and its supplementary information files].

## Declarations

### Ethics approval and consent to participate

Not applicable

### Author contributions

P-Y and X-L were responsible for the entire project and revised the draft. H.L-Z, S.S-H, and K.B-M performed the data extraction, statistical analysis, and interpreting the data. X.L, and S.S-H drafted the first version of the manuscript. All authors participated in the interpretation of the results and prepared the final version of the manuscript.

## Acknowledgments

None

## Consent for publication

Not applicable

